# Precision Mapping of Thalamic Deep Brain Stimulation Lead Positions Associated with the Microlesion Effect in Tourette Syndrome

**DOI:** 10.1101/2022.09.07.22279661

**Authors:** Takashi Morishita, Yuki Sakai, Hitoshi Iida, Saki Yoshimura, Shinsuke Fujioka, Kazunori Oda, Saori C. Tanaka, Hiroshi Abe

## Abstract

**Objectives:** The microlesion effect is refers to the improvement of clinical symptoms after deep brain stimulation (DBS) lead placement and is suggested to indicate optimal lead placement. This phenomenon is well known for movement disorders, such as essential tremors and Parkinson disease, but very few studies have reported its implications in neuropsychiatric disorders. Our aim was to evaluate the magnitude of the microlesion effect in Tourette syndrome (TS) and the relationship between the microlesion effect and the anatomical location of implanted DBS leads.

**Methods:** This study included 6 consecutive patients who underwent DBS for severe TS. All patients were male and their mean age was 28.5 ± 10.5 years. All patients were videotaped at baseline and on postoperative day (POD) 7, and motor and phonic tic frequencies were recorded. We also analyzed the precision of lead placement in normalized brain space and evaluated the normative connectome associated with precise electrode positions for improvement of tics.

**Results:** The microlesion effect was observed as an improvement in tic symptoms in all patients. The median motor tic frequency was 20.2 tics/min (range, 9.7–60) at baseline and decreased to 3.2 tics/min (1.2–11.3) on POD 1 (z = −2.20, p = 0.028) and 5.7 tics/min (range, 1.9–16.6) on POD 7 (z = −2.20, p = 0.028). The median phonic tic frequency was 10.5 tics/min (range, 2.0−58.7) at baseline and decreased to 0.7 tics/min (range, 0−14.4) on POD 1 (z = −1.78, p = 0.075) and 2.25 tics/min (range, 1.3−13.7) on POD 7 (z = −1.57, p = 0.116). Image analyses revealed that the precise position of the electrode was directed toward the anteromedial centromedian nucleus. Normative connectome analysis demonstrated connections between improvement-related areas and wide areas of the prefrontal cortex.

**Conclusion:** This study shows that the microlesion effect may appear as an immediate improvement following DBS lead placement even in patients with TS. Our results support the existence of a “sweet spot” for tic suppression in patients with severe TS, and clinicians should pay attention to this phenomenon in the postoperative evaluation of the lead position.

## Introduction

Tourette syndrome (TS) is characterized by tic movements that rarely result in a debilitating state. Deep brain stimulation (DBS) may be indicated in patients with severe symptoms refractory to medical and behavioral therapies. Since the success of the first reported application of DBS to TS,^1^ the clinical efficacy of the method has been confirmed in many reports.^2-4^

Although the first DBS was delivered to the centromedian (CM) thalamic nucleus, other targets, including the anterior limb of the internal capsule and the globus pallidus interna, are suggested to be equally effective.^3,4^ However, the therapeutic effects differ among patients. Although lead misplacement has potentially been linked to DBS failure,^5^ very few reports mention the possibility of DBS failure for TS.^6^ Because identification of the structure of the thalamic target is usually difficult in images, determination of the lead misplacement in thalamic DBS for TS is challenging, unless the misplacement is obvious. A recent multicenter analysis reported the preferred anatomical locations for TS-DBS,^7^ although the ideal position of the DBS electrode remains unclear.

The microlesion effect has been reported as a clinical sign of optimal lead placement in DBS for essential tremor and Parkinson disease.^8,9^ Here, we further investigate a patient whom we previously studied, in which temporary tic suppression persisted for 1 postoperative week,^6^ in addition to the inclusion of new patients. We hypothesized that a “sweet spot” area is associated with the microlesion effect of lead placement and optimal electrical stimulation in patients undergoing thalamic DBS for severe TS. Thus, this study evaluated the relationship between the microlesion effect and the anatomical location of implanted DBS leads in these patients.

## Methods

### Study design

The clinical data of the patients were prospectively recorded, and their charts were reviewed to obtain detailed information. DBS lead position was identified in standard brain space. The study protocol was approved by the review board of our institute, Fukuoka University Medical Ethics Review Board (approval numbers: U02-02-001 and U22-02-012). Informed consent was obtained from all participants. The study was conducted according to the principles of the Declaration of Helsinki.

Consecutive patients undergoing CM thalamic DBS in our department from January 2020 to December 2021 were included in this study. Of these, 1 patient (Case 1) was included in our previous study.^6^ In our facility, DBS therapy is indicated for patients ≧ 12 years of age with severe medication-refractory symptoms. As previously described, a multidisciplinary evaluation was performed in all patients, as recommended by recent guidelines.^6,10^

The primary outcome measure was tic frequency evaluated in video recordings. We videotaped patients on postoperative days (PODs) 1 and 7 in the off-DBS condition to evaluate the microlesion effect and at 6 months of follow-up in the on- and off-DBS conditions. Study participants were hospitalized for 6 months of follow-up. Following videotape recording in the on-DBS condition and a 24-h washout period, the patients were videotaped again in the off-DBS condition. During each session, the patients were left alone in a quiet room with a video camera and videotaped for at least 5 min. Tic frequency was determined by a rater (S.Y.) who was blinded to the clinical status (i.e. DBS status) of the patients. We also evaluated clinical outcomes with the Yale Global Tic Severity Scale (YGTSS)^11^ both preoperatively and at 6 months after surgery. Additionally, adverse events were recorded. During the 6-month study period, the medications were unchanged, except in 1 patient (Case 6).

### Surgical procedure

Our DBS procedures have been reported in previous studies.^6,12,13^ High-resolution magnetic resonance imaging (MRI) was conducted using the same MRI scanner (Ingenia 1.5T, Philips, The Netherlands), and the MRI sequences comprised volumetric fast gray matter acquisition T1 inversion recovery and volumetric T1-weighted imaging (T1WI) with contrast. Stereotactic planning for DBS lead placement was performed using iPlan Stereotaxy stereotactic planning software (BrainLab, Germany). The MRI sequences were fused automatically, and the mid-commissural point was defined to anchor the Cartesian coordinate system. We initially set the tentative target at 4-mm posterior and 5-mm lateral to the mid-commissural point on the anterior commissure-posterior commissure plane. The trajectory was specifically chosen to ensure that the DBS lead would avoid the sulci, lateral ventricle, and vasculature, and in each patient, the target was modified according to the insertion angle.

In this study, simultaneous bilateral DBS lead implantation (model 3387, Medtronic, MN, USA) was conducted under C-arm guidance, and we implanted an implantable pulse generator (IPG) on the same day (Activa RC for Cases 1 and 2; Percept PC from Case 3 on; Medtronic). All patients underwent the lead implantation while asleep under general anesthesia or deep sedation and the IPG implantation under general anesthesia.

Computed tomography (CT) was used to identify the lead location, typically on POD 9, after resolution of the postoperative pneumocephalus. The CT image was fused with a preoperative scan to rule out lead misplacement. In cases where lead misplacement was diagnosed, additional surgery was performed during the same admission period.

### DBS programming

Electrical stimulation was initiated 2 weeks after DBS surgery to activate the second-most ventral contacts bilaterally, with the following parameters: intensity, 2.0 mA; pulse width, 60; and frequency, 130 Hz. Each patient was instructed to attend our DBS clinic once a month for 6 months of follow-up, and the stimulation intensity was gradually increased. At each visit, any new programming parameters were saved as a new group setting, which enabled patients to return to the previous setting if any tic aggravation or mood changes developed. If the stimulation parameter reached the maximum intensity within the therapeutic window in a single monopolar setting, the adjacent contact was activated to apply interleaving or multiple monopolar stimuli.

### Image processing

This image processing procedure was reported in our previous studies.^6,25^ We used Lead-DBS (http://www.lead-dbs.org/) to preprocess imaging data.^14^ We coregistered images inclusive of postoperative CT and preoperative MR images linearly to the preoperative T1-weighted images using SPM software (http://www.lead-dbs.org/). To minimize the nonlinear bias associated with DBS surgery, the registration between preoperative T1WI and postoperative CT was refined using linear registration within the target region of interest in the subcortex.^15^ We normalized all data to standard Montreal Neurological Institute space. The procedure was conducted using whole-brain nonlinear SyN registration in Advanced Normalization Tools (ANTs, http://stnava.github.io/ANTs/)^16^ with the “effective (low variance)” setting and with subcortical refinement carried out in Lead-DBS.^14^ We automatically pre-reconstructed electrode trajectories and contacts using the PaCER algorithm^17^ and manually adjusted them with Lead-DBS. Subject-independent 3D atlases of the thalamic nuclei were applied to evaluate the lead trajectory and contact positions.^18^

### Creation of the presumed microlesion area

Precise identification of microlesion areas is difficult, so we assumed that microlesion areas would form around the electrode trajectories and create a presumed microlesion area. All of the following procedures were performed in a voxel space of 0.5 × 0.5 × 0.5 mm.

First, we delineated the line between the most ventral and dorsal contacts identified using the PaCER algorithm. Because we needed to represent this line in voxel space, we used the line algorithm of 3D Bresenham (Figure 1A). We then performed a 3-voxel level dilation of this line using the 3dmask_tool of AFNI^19^ to create the surrounding presumed microlesion area, considering the microhemorrhage and edema (Figure 1B). An example of a presumed microlesion area in standard brain space is shown in Figure 2C. Because the volume of data was limited, we used ANTs to nonlinearly transform the presumed microlesion areas in the right hemisphere into those in the left hemisphere. All presumed microlesion areas were pooled across the hemispheres as left-sided areas.

**Figure 1.**
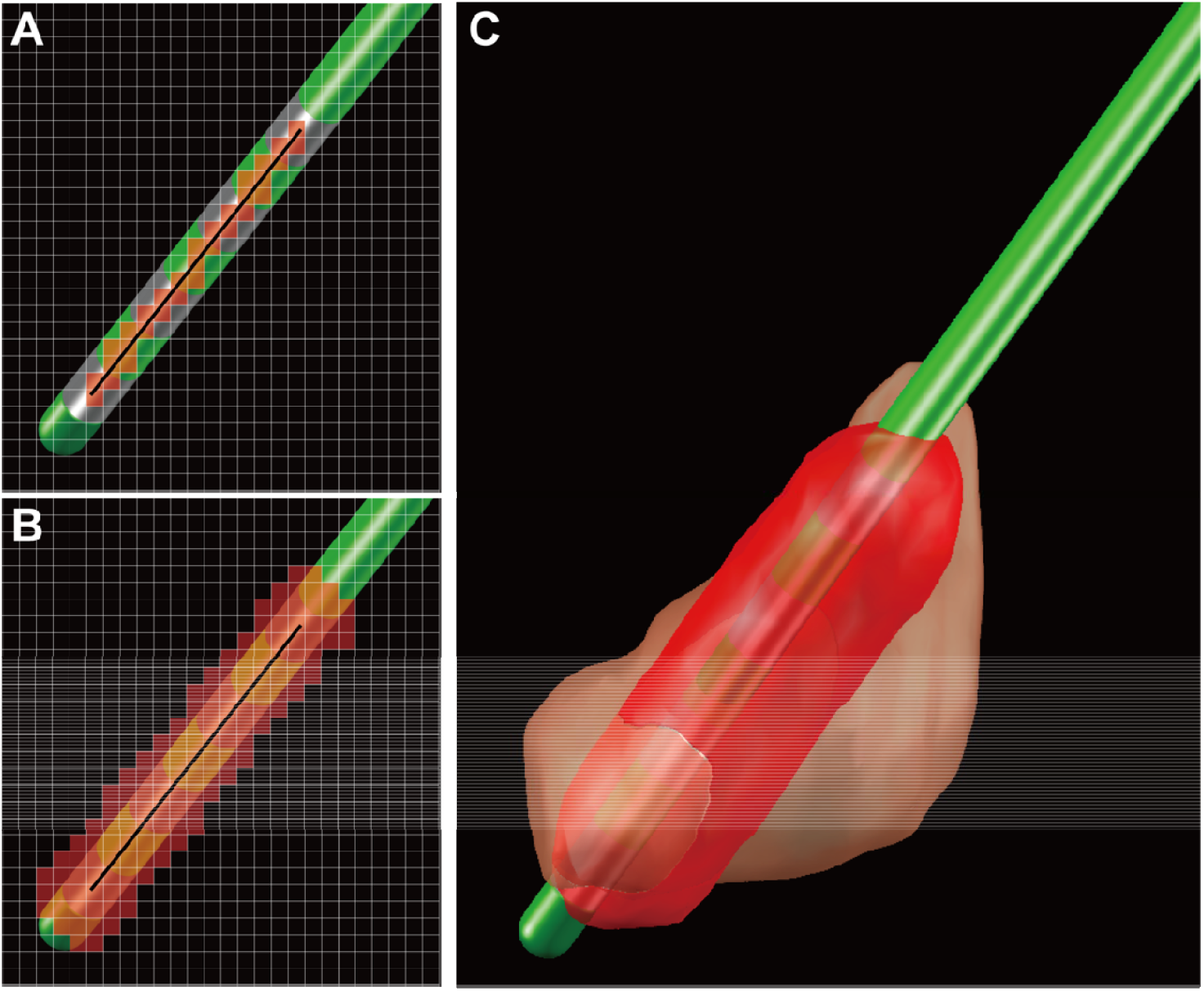
Creation of a presumed microlesion area along the deep brain stimulation (DBS) electrode. (A) Line between the most ventral and dorsal contacts. The black line and the red area represent the center line of the electrode and the line in the voxel space using the line algorithm of 3D Bresenham, respectively. The grid represents a voxel space of 0.5 × 0.5 mm. (B) The 3-voxel level dilation of the line in Figure 2A was created using the 3dmask_tool of AFNI to create the surrounding presumed microlesion area. The grid represents a voxel space of 0.5 × 0.5 mm. (C) An example of a presumed microlesion area in standard brain space. The peach-colored region is the CM nucleus.

**Figure 2.**
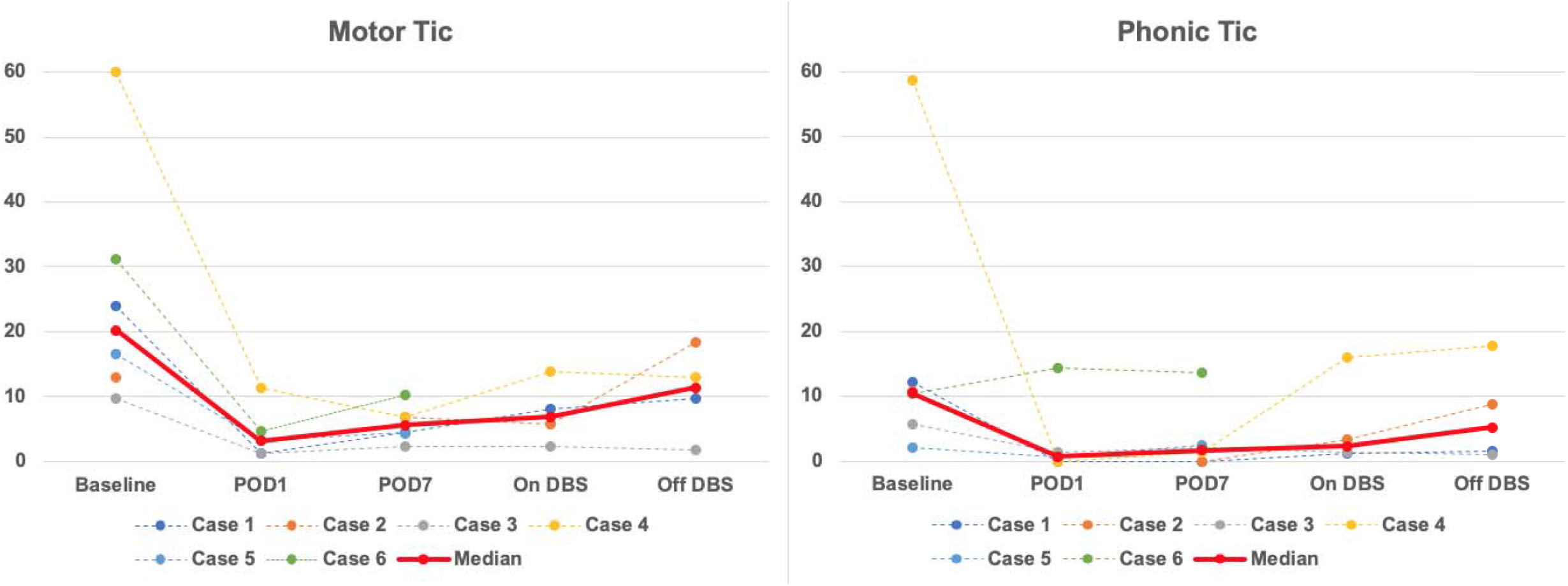
Chronological changes in video-rated tic frequency (tics/min). The tic frequencies on postoperative days (PODs) 1 and 7 were evaluated in the off-DBS condition, whereas the on- and off-DBS conditions were evaluated at the 6-month follow-up. In this figure, the POD numbers were counted after the revision surgery of Case 4.

To precisely evaluate the spatial overlap of the microlesion areas between patients in standard brain space, we assessed the percentage overlap of all microlesion areas. To further evaluate the relationship between microlesion area and symptom improvement, we calculated the rate of symptom improvement relative to the preoperative severity. We calculated the improvement rate separately for motor and phonic tics and used the mean value. The median value among patients was assigned to each voxel in overlapping areas. When a presumed microlesion area of an electrode did not contain a voxel, its voxel improvement rate was regarded as 0. We defined the depicted region as an improvement-related area.

### Normative connectome analysis

To characterize the brain connectome from the microlesion area and reveal how microlesions affect the entire brain, we applied the population-averaged atlas of the macroscale human structural connectome derived from diffusion-weighted imaging data (N = 842, Human Connectome Project).^20^ Because we considered the improvement-related area to be the core region of microlesion effects, we evaluated the fiber tracts passing through it. In addition, we assessed how many structural fibers are connected to each brain area determined by the Harvard-Oxford cortical/subcortical atlases^21^ in conjunction with the AAL atlas cerebellum.^22^ All small AAL-defined cerebellar parcels were integrated into a single binarized parcel in a manner similar to that in our previous report.^6^

### Statistical analysis

We used the Wilcoxon signed-rank test to compare the pre- and post-DBS clinical scores of the YGTSS and tic frequencies. Statistical analyses were performed using SPSS version 21.0 (IBM Corp., Armonk, NY, USA), and p < 0.05 indicates statistical significance.

## Results

### Clinical outcomes

This study included 6 participants. Median age at onset of TS and surgery was 8 years (range, 3−9) and 25 years (range, 18−47), respectively. All 6 participants were male. Video evaluation was performed during hospitalization immediately after DBS surgery in all 6 participants, but just 4 of the 6 participants (Cases 1–4) completed the planned 6-month follow-up. One patient was lost to follow-up and another could not complete the 6-month video rating. The median YGTSS motor and phonic scores improved from 23.5 (range, 22−44) and 19.5 (range, 14−21) at baseline to 7.0 (range, 0−9) (z = −2.20, p = 0.028) and 7.0 (range, 7−12) (z = −2.20, p = 0.028) at the final follow-up, respectively. The median YGTSS impairment score improved from 50 (range, 40−50) to 30 (range, 10−30) (z = −2.20, p = 0.028). The clinical outcomes are summarized in Table 1.

**Table 1.** Patient demographics and YGTSS scores. **Childhood: age 3-12; Adolescence: age 13-19, Young adult: age 20-39, Adult: age >39.**

| Case | Sex | Age of Onset | Age at Surgery | YGTSS Severity Motor |  | YGTSS Severity Phonic |  | YGTSS Impairment |  |
| --- | --- | --- | --- | --- | --- | --- | --- | --- | --- |
|  |  |  |  | Baseline | 6 Mos | Baseline | 6 Mos | Baseline | 6 Mos |
| 1 | Male | Childhood | Adolescence | 23 | 5 | 20 | 7 | 40 | 10 |
| 2 | Male | Childhood | Young adult | 44 | 9 | 21 | 12 | 50 | 30 |
| 3 | Male | Childhood | Young adult | 22 | 7 | 15 | 7 | 50 | 30 |
| 4 | Male | Childhood | Young adult | 22 | 0 | 19 | 8 | 40 | 10 |
| 5 | Male | Childhood | Young adult | 25 | NA | 14 | NA | 50 | NA |
| 6 | Male | Childhood | Adult | 24 | 7 | 21 | 7 | 50 | 30 |

The microlesion effect was observed as an improvement in tic frequency in all patients. The median motor tic frequency was 20.2 tics/min (range, 9.7−60) at baseline and decreased to 3.2 tics/min (range, 1.2−11.3) on POD 1 (z = −2.20, p = 0.028) and 5.7 tics/min (range, 1.9−16.6) on POD 7 (z = −2.20, p = 0.028). However, no improvement in phonic tics was observed during the immediate postoperative period in either of the 2 patients who did not complete the full 6-month follow-up (Cases 5 and 6). The median phonic tic frequency was 10.5 tics/min (range, 2.0−58.7) at baseline and decreased to 0.7 tics/min (range, 0−14.4) on POD 1 (z = −1.78, p = 0.075) and 2.25 tics/min (range, 1.3−13.7) on POD 7 (z = −1.57, p = 0.116). At the 6-month evaluation, the motor and phonic tic frequencies in the on-DBS condition were 6.9 tics/min (range, 2.4−13.8) and 2.3 tics/min (range, 1.0−15.9), respectively. These scores were significantly lower than those at baseline (motor: z = −2.20, p = 0.028; phonic: z = −2.20, p = 0.028). In addition, the motor and phonic tic frequencies in the off-DBS condition were 11.4 tics/min (range, 1.8−18.4) and 5.2 tics/min (range, 1.0−17.8), respectively. These scores were also significantly lower than those at baseline (motor: z = −1.99, p = 0.046; phonic: z = −1.99, p = 0.046). The chronological changes are shown in Figure 2. The stimulation parameters at the 6-month follow-up are summarized in Table 2.

**Table 2.**
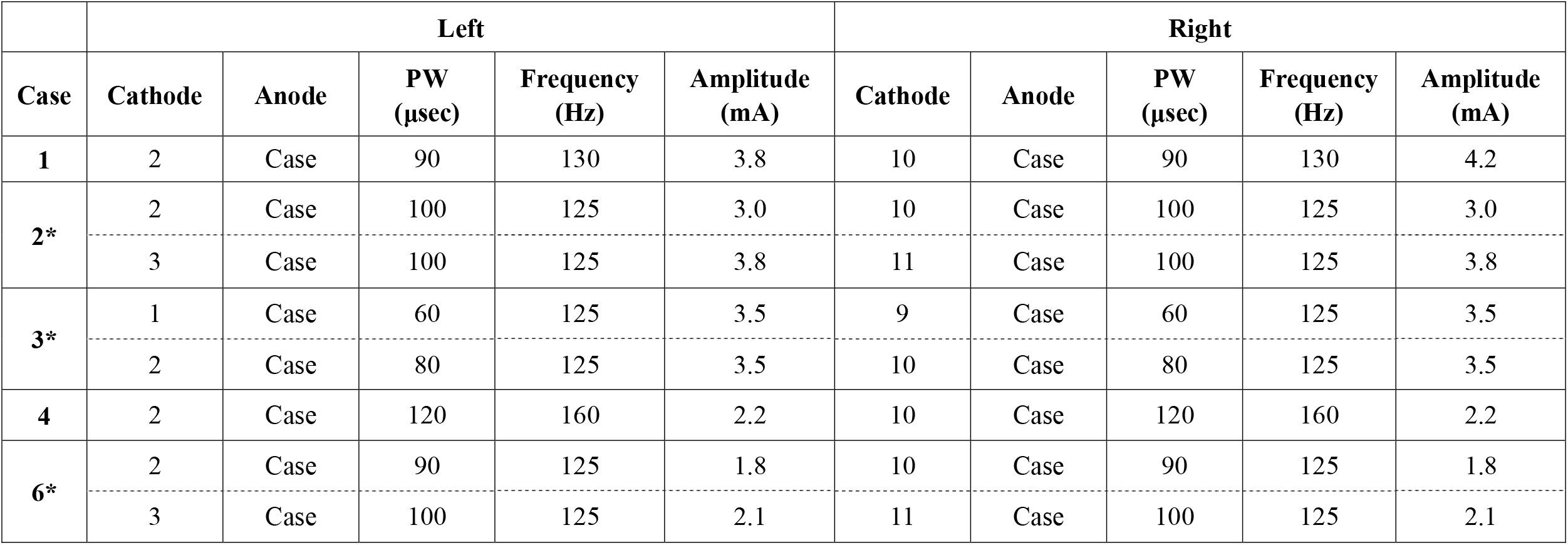
The stimulation parameters at six-month follow up. *Interleaving stimulation settings were applied. Abbreviation. PW = pulse width

### Adverse events

Surgical adverse events included wound dehiscence treated with surgical debridement (Case 3). Stimulation-induced depression was also reported in the same patient, but this adverse event was promptly resolved by adjusting the stimulation parameters. Lead misplacement on the left was diagnosed in Case 4 on POD 9; therefore, the patient underwent revision surgery on POD 12 before discharge. We excluded misplaced leads from the imaging analysis. Another patient (Case 6) experienced intracerebral hemorrhage along the left DBS electrode due to severe self-injurious tics. To prevent expansion of the hemorrhage, we had to sedate the patient for several days and increase the medication and stimulation intensity to control the violent tics.

### Stereotactic precision and connectome images

The DBS lead locations in normalized brain space are shown in Figure 3. In our cohort, precision analysis showed that the presumed microlesion areas for 8 of 12 DBS electrodes (75%) overlapped along the electrode trajectory through the ventromedial area of the ventral lateral nucleus to the anteromedial area of the CM nucleus (Figure 4A). Image analysis showed that the precise electrode position was centered on the anteromedial CM nucleus (Figure 4B). Normative connectome analysis showed connections between the improvement-related area and wide areas of the prefrontal cortices and globus pallidus (Figure 5).

**Figure 3.**
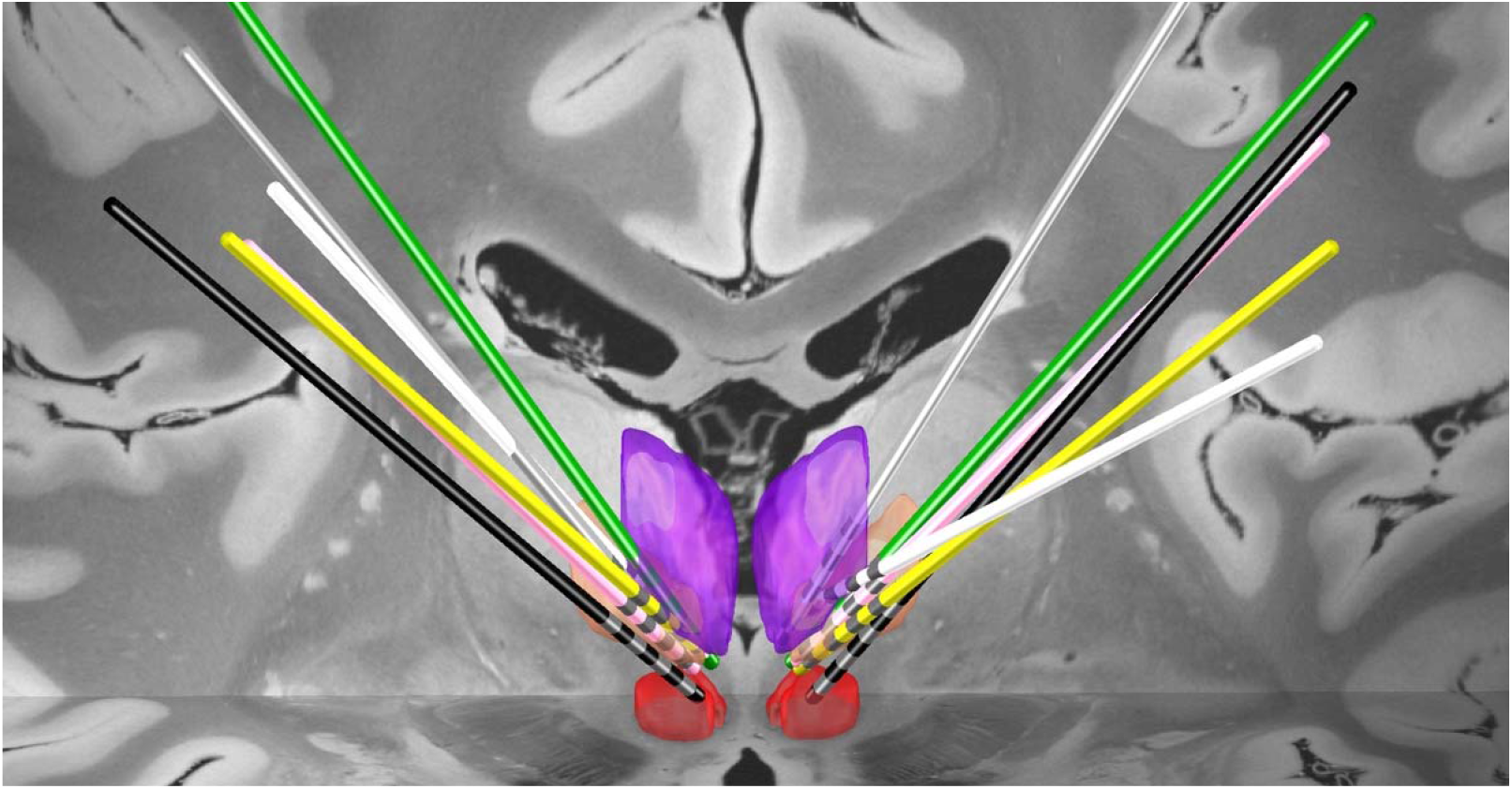
Lead electrode placement in standard brain space. Lead electrodes in the CM nucleus (peach), red nucleus (red), and mediodorsal nucleus (purple). The lead electrodes of each patient are displayed in different colors (Case 1, green; Case 2, pink; Case 3, white; Case 4, black; Case 5, yellow; Case 6, gray).

**Figure 4.**
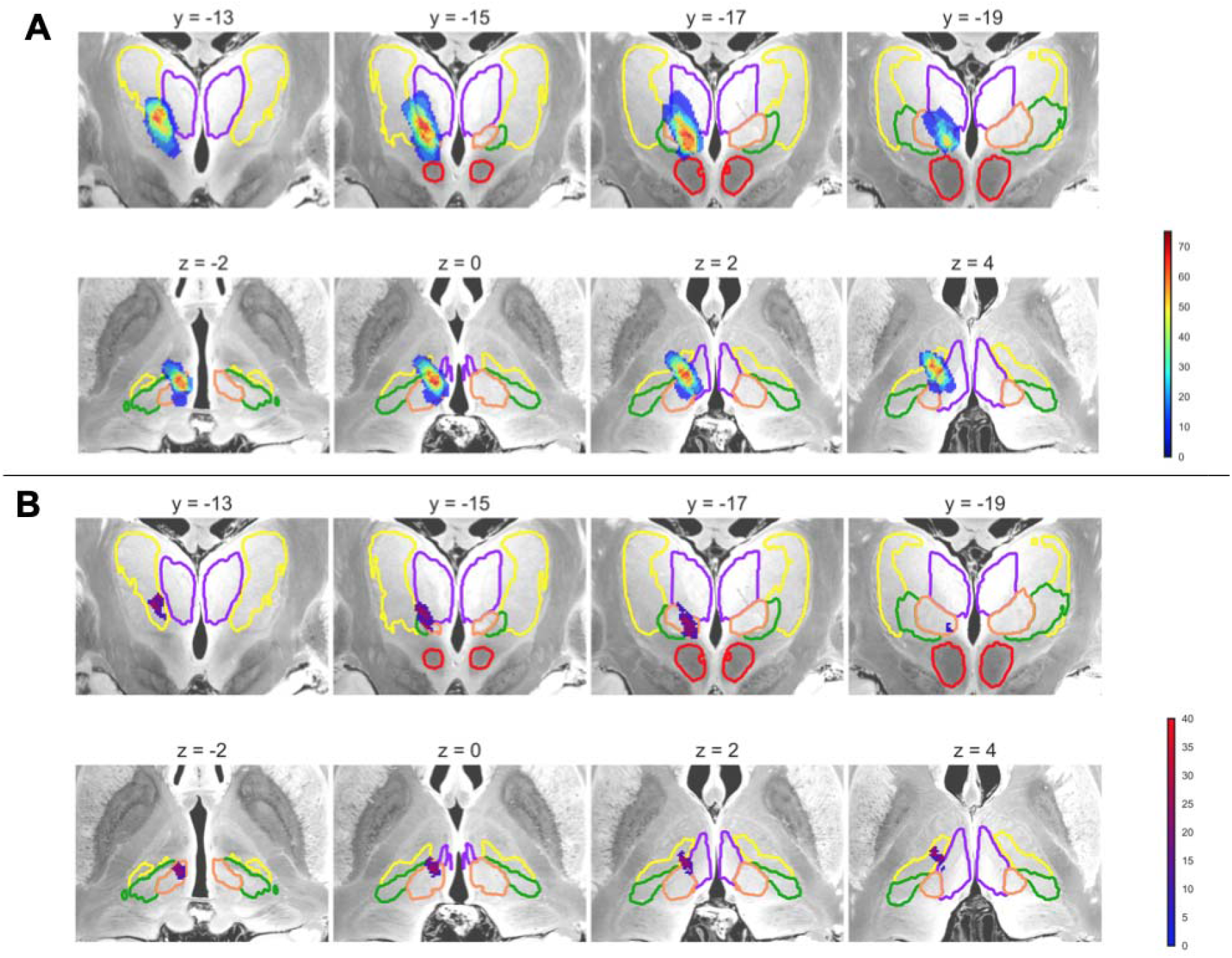
Precision mapping of implanted deep brain stimulation electrodes in an atlas. The upper and lower panels are the coronal and axial planes, respectively. Brain areas surrounded by a colored line represent the thalamic nuclei (CM nucleus, peach; red nucleus, red; mediodorsal nucleus, purple). The numbers above each panel are the Montreal Neurological Institute coordinates. (A) Heat map of the microlesion area overlaps. The color bar represents the percentage of overlap among patients. (B) Improvement-related area. The color bar represents the percentage improvement in tic symptoms. See Supplementary Movie 1 for a more detailed position compared to thalamic nuclei.

**Figure 5.**
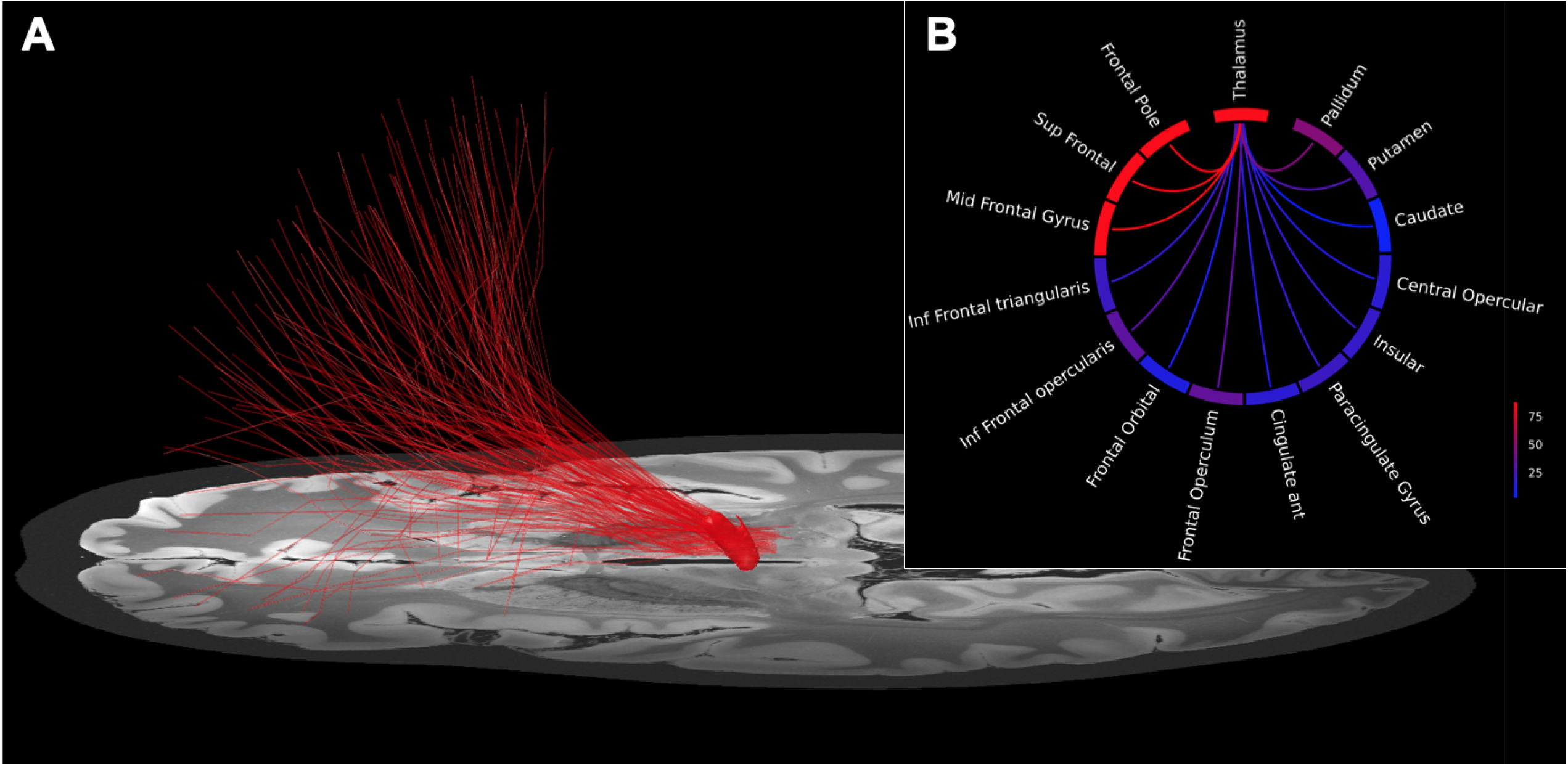
Normative connectome associated with symptom improvement-related areas. (A) Fiber tracts passing through the improvement-related area. (B) Circular plot of the number of structural fibers passing through each brain region. The color bar represents the number of structural fibers.

## Discussion

This study investigated microlesion effects in 6 cases of medication-refractory TS and the associated thalamic region. Because the microlesion effect has been reported to be a predictive factor for successful DBS lead placement in Parkinson disease and essential tremor, we hypothesized that this phenomenon could also occur in thalamic DBS for severe TS. Based on our findings, we recommend that clinicians be aware that microlesion effects may potentially alter stimulation effects and be cautious about interpreting the clinical findings in programming sessions at the early postoperative stage. In cases where a strong microlesion effect is observed immediately following DBS surgery, DBS effects may appear to be lost over time if the stimulation parameters are not appropriate. Ideally, stimulation parameters should be programmed to be minimal and focus on the sweet spot to avoid stimulation-induced adverse effects.

The microlesion effect is considered to result from edema, microhemorrhage, and the disruption of cells and/or fibers along the trajectory of DBS electrodes. The microlesion effect may even persist 6 months after surgery^8,9^ and, in some cases, symptoms may be completely abolished without stimulation.^24^ The long-lasting microlesion effect may partly explain why the tic frequency in the off-DBS condition in our cases was lower than the baseline level even after 24 h of stimulation washout (Figure 2), although it is possible that neuroplasticity may have been induced by chronic electrical stimulation.

In our previous study, we applied the volume of tissue activated (VTA) approach and the related normative connectome analyses and suggested the existence of a sweet spot for tic suppression in patients with severe TS.^6^ Additionally, we reported that the sweet spot could be the anterior border of the CM nucleus and the medial area of the ventral lateral nucleus as the optimal stimulation area in other studies.^2,25^ The present study further supports this possibility (Figure 4). Furthermore, the normative connectome associated with the microlesion effect has components common to the connectome associated with the VTA of the therapeutic stimulation area; therefore, we consider that the presumed microlesion area in this study may include the sweet spot.

Normative connectome analysis has been increasingly used as a reliable research tool in DBS. As shown in the present and previous studies,^6,25^ the normative connectome supports the interpretation of clinical effects and potentially guides DBS programming to induce therapeutic effects. Obtaining high-quality diffusion tensor images preoperatively is challenging because of the acquisition time and signal-to-noise ratio. However, high-quality images are used in normative connectome analysis. Furthermore, a recent study showed similar quality in analytical data between normative and patient-specific connectomes.^26^ Therefore, we consider that the application of normative connectome analysis in this study is a reasonable option to support the findings of precision mapping.

It should also be noted that DBS surgery is usually performed bilaterally for neuropsychiatric diseases. However, there is a controversy regarding laterality.^27,28^ Interestingly, one patient (Case 4) in our cohort showed clinical improvement immediately after the first surgery, although the DBS electrode was misplaced on one side. Additionally, the patient showed further improvement following revision surgery. In this context, there is a possibility that unilateral or staged surgery may be beneficial in selected patients, but no conclusion can be reached here. In addition, we reported another case of lead misplacement in a previous study,^6^ and this issue could raise concern regarding the difficulty of precise lead placement in the CM thalamic DBS procedure.

Although our study suggests that the microlesion effect may help to predict favorable outcomes of DBS therapy in severe medication-refractory TS, it has several limitations. Only 6 patients were enrolled in this study. Although the microlesion effect persisted at the 6-month follow-up, other possible mechanisms, such as neuroplastic changes, have not been investigated. In terms of the imaging study, we assumed and defined the microlesion area as a constant volume among patients; however, this may not be accurate because of the heterogeneity of the density of brain tissue or other biological factors. Because severe, medication-refractory TS is a rare disorder, multicenter studies with larger numbers of patients are needed to confirm our findings.^4,7,29^

## Conclusion

This study showed that the microlesion effect may appear as an immediate improvement following DBS lead placement, even in TS, and this phenomenon may help to predict a favorable clinical outcome, as reported in Parkinson disease and essential tremor. Our results support the existence of sweet spots for tic suppression in patients with severe TS. Clinicians should pay attention to programming sessions in the clinical setting. Further studies with greater patient numbers are required.

## Data Availability

All data produced in the present study are available upon reasonable request to the authors.

## Abbreviations

ANTs: Advanced Normalization Tools;
CM: centromedian;
CT: computed tomography;
DBS: deep brain stimulation;
IPG: implantable pulse generator;
MRI: magnetic resonance imaging;
TS: Tourette syndrome;
VTA: volume of tissue activated;
YGTSS: Yale Global Tic Severity Scale

## Acknowledgments

None.

## Author Contributions

TM and YS contributed to the conceptualization, data collection and analysis, and manuscript writing. HI, SY, and SF contributed to the data gathering and review of the manuscript. KO contributed to the statistical analysis. SCT and HA supervised the study and reviewed the manuscript.

